# Time to embrace the molecular approach for respiratory pathogen screening

**DOI:** 10.1101/2025.08.08.25332920

**Authors:** Amélia Rodrigues, Maria Eduarda Cazelli, Margarida Eulálio, Sónia Ferreira, Ana Sousa

## Abstract

The current gold standard for clinical pathogen identification relies on a combination of culture-based methods, target-specific molecular techniques, and antigen-detection assays^1^. Known limitations of the first method include the requirement for viable and culturable microorganisms, while the second approach is constrained to a limited number of predefined targets. These constraints reduce their diagnostic success, as not all pathogens are culturable, easily isolated from commensal microbiota, or represented in the selected detection panels. While culture-based methods are widely recognized for their limited sensitivity, updated quantifications of their actual performance in real-world clinical settings are scarce.

To address this gap, we conducted a retrospective survey of all lower respiratory tract (LRT) samples processed by either culture or target-based molecular methods over a five-year period at a Portuguese hospital centre, providing a quantitative assessment of current diagnostic performance.

## Results and Discussion

The study was approved by the Health Ethics Committee of the Unidade Local de Saúde Região de Aveiro (ULSRA) (N/Ref^a^42-032021, 11/05/2022). The requirement to obtain informed written consent from each individual was waived as the study was limited to the data obtained from ULSRA database, without the need to identify patients.

Representative samples were obtained from emergency, inpatient, and outpatient consultations at ULSRA between January 2017 and December 2022, with the exception of *Mycobacterium tuberculosis*, for which data collection began in January 2018. Sars-Cov2 virus data was not included in the analysis.

Sputum, endotracheal aspirate (ETA), bronchoalveolar aspirate (BAA) and lavage (BAL) were analysed at the pathology service and processed according to the clinician’s request, which may include cultural analysis, respiratory PCR panel (bacterial and/or viral PCR panel) and *M. tuberculosis* (see supplements for methodological details).

Excluding *M. tuberculosis*, the final study population comprised 7455 samples tested using culture-based methods, and 825 samples analysed by molecular approach. For *M. tuberculosis* detection a total of 11361 samples were tested. Demographic characteristics for different samples are presented in Table 1. From the 7455 samples tested using a culture-based approach, only 1820 (24%) resulted in a positive identification (Fig 1A), which is similar to previous reports from other healthcare institutions ^2^. The product with the highest rate of positive identifications was ETA, with 40%, and the lowest was BAL, which led to 15% positive identifications.

**Table 1:** Number and type of biological samples (sputum, endotracheal aspirate, bronchoalveolar aspirate and bronchoalveolar lavage) used for diagnosis of LRT infections in a 5 year period in a Portuguese ULS. The methodological approach: culture-based, bacteriological/viral PCR panel and *Mycobacterium tuberculosis* detection is also indicated. N.A. indicate tests that are not routinely performed at the ULS.

|  | SPUTUM |  |  | Endotracheal aspirate (ETA) |  |  | Bronchoalveolar aspirate (BAA) |  |  | Bronchoalveolar lavage (BAL) |  |  |
| --- | --- | --- | --- | --- | --- | --- | --- | --- | --- | --- | --- | --- |
|  | n | % | Age (years)<br>mean±SD | n | % | Age (years)<br>mean±SD | n | % | Age (years)<br>mean±SD | n | % | Age (years)<br>mean±SD |
| **Culture-based** |  |  |  |  |  |  |  |  |  |  |  |  |
| Female | 1459 | 40,2% | 66,7±18 | 296 | 30,6% | 61,1±16 | 747 | 38,5% | 63±16 | 405 | 43,3% | 61,3±16 |
| Male | 2171 | 59,8% | 66,1±17 | 671 | 69,4% | 61±14 | 1194 | 61,5% | 64,3±15 | 512 | 56,7% | 63,6±16 |
| **Total** | **3630** |  |  | **967** |  |  | **1941** |  |  | **917** |  |  |
| **Bacteriologic PCR panel** |  |  |  |  |  |  |  |  |  |  |  |  |
| Female | N.A. |  |  | N.A. |  |  | N.A. |  |  | 261 | 44,4% | 59,7±18 |
| Male | N.A. |  |  | N.A. |  |  | N.A. |  |  | 313 | 55,6% | 62,3±17 |
| **Total** | N.A. |  |  | N.A. |  |  | N.A. |  |  | **574** |  |  |
| **Viral PCR panel** |  |  |  |  |  |  |  |  |  |  |  |  |
| Female | N.A. |  |  | N.A. |  |  | N.A. |  |  | 110 | 42,9% | 58,1±18 |
| Male | N.A. |  |  | N.A. |  |  | N.A. |  |  | 141 | 57,1% | 60,2±20 |
| **Total** | N.A. |  |  | N.A. |  |  | N.A. |  |  | **251** |  |  |
| ***M. tuberculosis*** |  |  |  |  |  |  |  |  |  |  |  |  |
| Ziehl-Nielsen Stain |  |  |  | N.A. |  |  |  |  |  |  |  |  |
| Female | 809 | 34,9% | 62,5±18 | N.A. |  |  | 570 | 39,2% | 63,6±16 | 371 | 42,9% | 61,6±16 |
| Male | 1569 | 65,1% | 60,1±18 | N.A. |  |  | 860 | 60,8% | 65,3±15 | 479 | 63,5% | 63,5±16 |
| **Total** | **2378** |  |  | N.A. |  |  | **1430** |  |  | **850** |  |  |
| **Culture** |  |  |  |  |  |  |  |  |  |  |  |  |
| Female | 769 | 38,4% | 64±17 | N.A. |  |  | 509 | 40,0% | 63,5±16 | 373 | 43,6% | 61,5±16 |
| Male | 1234 | 61,6% | 60,4±18 | N.A. |  |  | 778 | 60,6% | 65,6±14 | 465 | 56,4% | 63,7±15 |
| **Total** | **2003** |  |  | N.A. |  |  | **1287** |  |  | **838** |  |  |
| **PCR** |  |  |  |  |  |  |  |  |  |  |  |  |
| Female | 475 | 45,5% | 62,6±17 | N.A. |  |  | 342 | 39,3% | 62,2±16 | 274 | 41,7% | 61,7±16 |
| Male | 601 | 54,5% | 59,3±18 | N.A. |  |  | 514 | 60,7% | 63,9±16 | 369 | 58,3% | 63±16 |
| **Total** | **1076** |  |  | N.A. |  |  | **856** |  |  | **643** |  |  |

**Figure 1.**
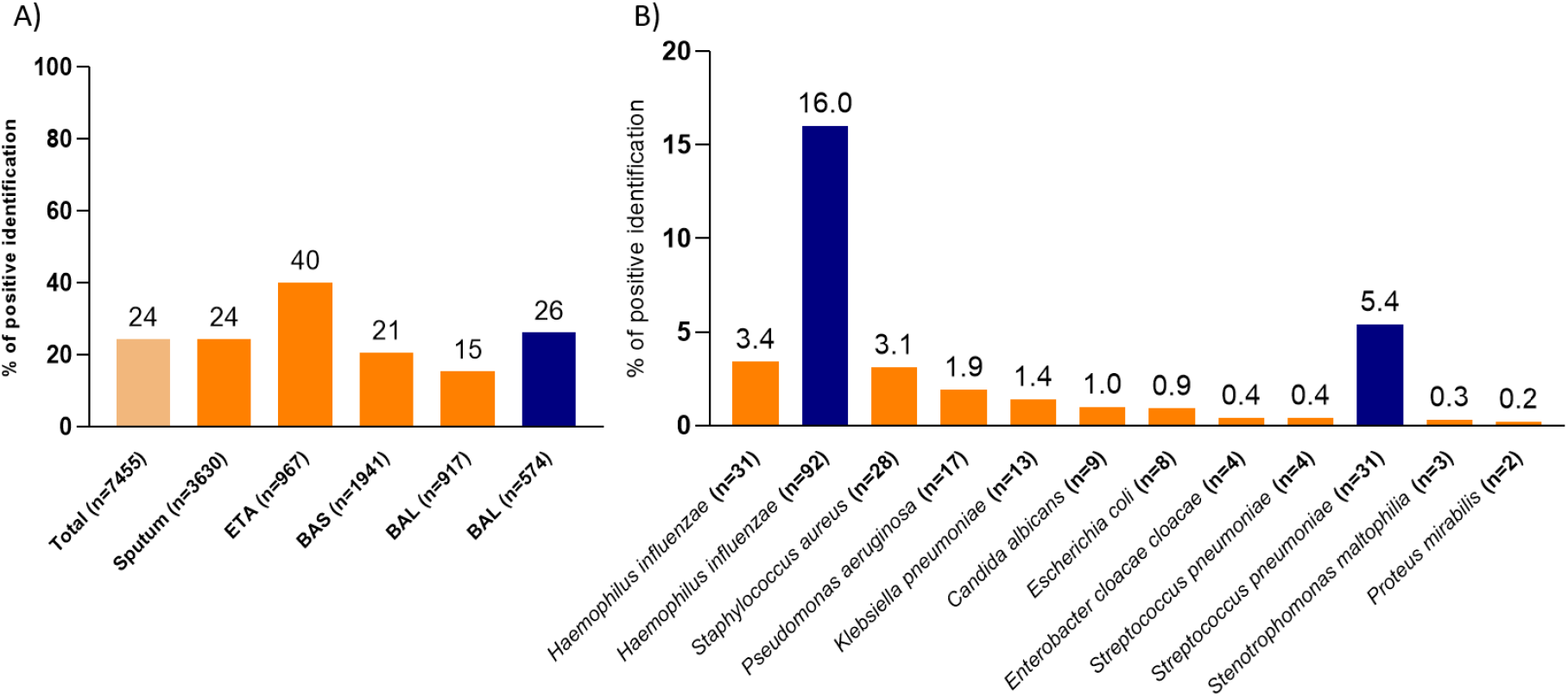
Percentage of bacterial/fungal positive identifications by culture-based methods (in orange) and by target molecular approach (in blue), using different types of biological samples (sputum, endotracheal aspirate (ETA), bronchoalveolar aspirate (BAA) and bronchoalveolar lavage (BAL)). Numbers of tested samples are indicated below each column. A) % of positive pathogen identifications indicated per sample type. B) % of positive pathogen identifications indicated per bacterial/fungal species. Only the top 10 most frequent bacterial species are shown.

In contrast, the molecular approach for bacterial detection in BAL led to 10% higher positive identifications (26%) (Fig 1A), despite being limited to a panel of only six bacterial species (including four low frequent pathogens) (Fig 1B). Specifically, *H. influenzae* was detected in only 3.4% by culture-based and in 16% of samples by the molecular approach, *i*.*e*., only one in four samples detected by molecular methods could also be detected by culture. For *S. pneumoniae*, the most frequent cause of community-acquired pneumonia ^3^, the rate of positive identifications was even lower, with the culture-based positive detection in 0.4% whereas the molecular methods could detect it in 5.4% of the samples (one in sixteen).

Another important difference between the two methods is the ability to detect co-infections, very rarely (less than 3%) the cultural-based approach identified more than one pathogen per sample, whereas the molecular approach identified the co-occurrence of two or more potential pathogens in about 14% of positive samples, in particular *S. pneumoniae* and *H. influenzae* co-occurred in 10% of these cases (Table S1).

These findings highlight the high false-negative rate associated with culture-based methods and underscore their limited resolution in capturing the complexity of respiratory infections—particularly in cases of co-infection and of fastidious bacteria such as *S. pneumoniae*, whose growth is often masked by commensal microbiota.

Considering all sample types, nine bacterial species and one fungus accounted for 86% of all positive identifications (Table S2), with 54 additional bacterial and fungal species comprising the other 14% positive identifications.

This illustrates, the large diversity of bacterial and fungal species that can be involved in LRT infections and highlights the importance of unbiased methods of diagnosis, in contrast with the target approaches that only capture a very small portion of this diversity.

The presence of *M. tuberculosis* can be assessed by several methods: 1) direct inspection of Ziehl– Neelsen staining, 2) culturing and by 3) molecular approach (target PCR). As expected, the direct inspection provided the lowest positive identifications of *M. tuberculosis*, since it heavily relies on the skill level of the person performing the analysis. The molecular approach yielded either a higher rate of positive identifications (15.3% vs. 3.2% by culture-based methods; Fig. S1) or comparable results. Moreover, it significantly reduced turnaround time, delivering results in approximately six hours compared to 3-8 weeks required for culture-based diagnostics. Nonetheless, culture-based methods will likely remain essential for monitoring patients undergoing antibiotic therapy, particularly in cases where confirming the viability of *M. tuberculosis* is critical ^4^.

During the considered period, viral detection in LRT samples was performed in 251 BAL samples. 12% of the samples provided a positive identification, with the top three viruses being Influenzae A or B (∼6%) and Rhinovirus (2%) (Table S3). Among viral infections, two out of 30 positive samples were co-infections.

A major limitation of current diagnostic methods is their inability to detect bacterial/fungal–viral co-infections in an unbiased manner within a single test, leading to these cases being rarely identified in routine practice. To better understand their frequency, we surveyed instances of positive viral diagnoses that coincided with bacterial infections and found that co-infections occurred in over 60% of cases. Such situations, might benefit from complementary antibacterial treatment, highlighting the need for diagnostic approaches capable of simultaneously detecting both viral and bacterial pathogens ^5^.

From our point of view, the conducted survey shows that it is essential to prepare the next stage, where metagenomics, offering a faster, broader, and more sensitive alternative for pathogen detection, could be an invaluable alternative which is just around the corner ^6^.

### Declaration of Generative AI and AI-assisted technologies in the writing process

During the preparation of this work the authors used ChatGPT-4o to improve readability and language. After using this tool/service, the authors reviewed and edited the content as needed and take full responsibility for the content of the publication.

## Supporting information

Manuscript

## Data Availability

All data produced in the present study are available upon reasonable request to the authors

