## Supplementary material for "Time to embrace the molecular approach for respiratory pathogen screening": Manuscript

### Methods

#### Culture-based identification of etiologic agents of infection

Samples used were: sputum, endotracheal aspirate (ETA), bronchoalveolar aspirate (BAS) and lavage (BAL). These were previously analysed at the ULSRA pathology service and processed according to the clinician's request, which may include cultural analysis, respiratory PCR panel (bacterial and/or viral PCR panel) and *Mycobacterium tuberculosis* (methodologic approach according to the medical request, see diagram below).

For bacterial search, samples were cultured on commercial laboratory-defined media: blood agar, chocolate agar and MacConkey agar. If mycological research was requested, culture in Sabouraud agar was also done. Cultures were incubated at 35ºC±2ºC, for at least 48 hours, unless identification could be obtained at 24 hours. Gram and Ziehl–Neelsen staining were also performed. Sputum Gram stains were screened according to the Murray-Washington (1975) criteria [1]. Sputum samples should only be considered for culture if >25 polymorphonuclear leukocytes and <10 squamous epithelial cells per 100× power field are found, with two exceptions: immunocompromised or neutropenic patients.

The infection agents were identified by matrix-assisted laser desorption/ionisation time-of-flight (MALDI-TOF) mass spectrometry (Bruker Portugal). The optochin susceptibility test was used to identify *Streptococcus pneumoniae*.

A negative result for identification was scored when: (1) no growth was obtained, (2) visible growth was obtained but no microorganism could be isolated (polymicrobial results without predominance) or (3) one or more microorganisms were isolated but these were not considered as causative agents of respiratory infection.

**<u>Respiratory samples</u>**

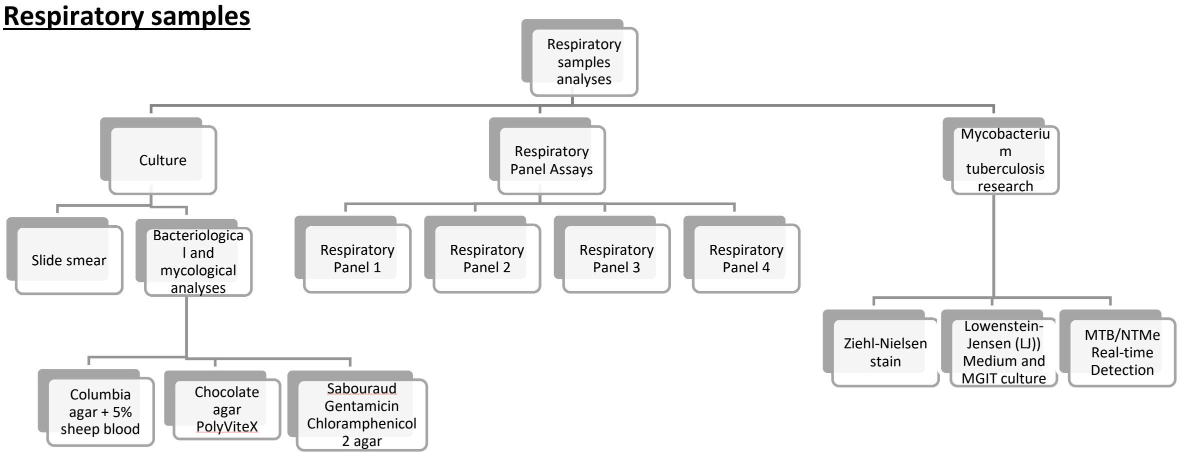

***Mycobacterium tuberculosis***

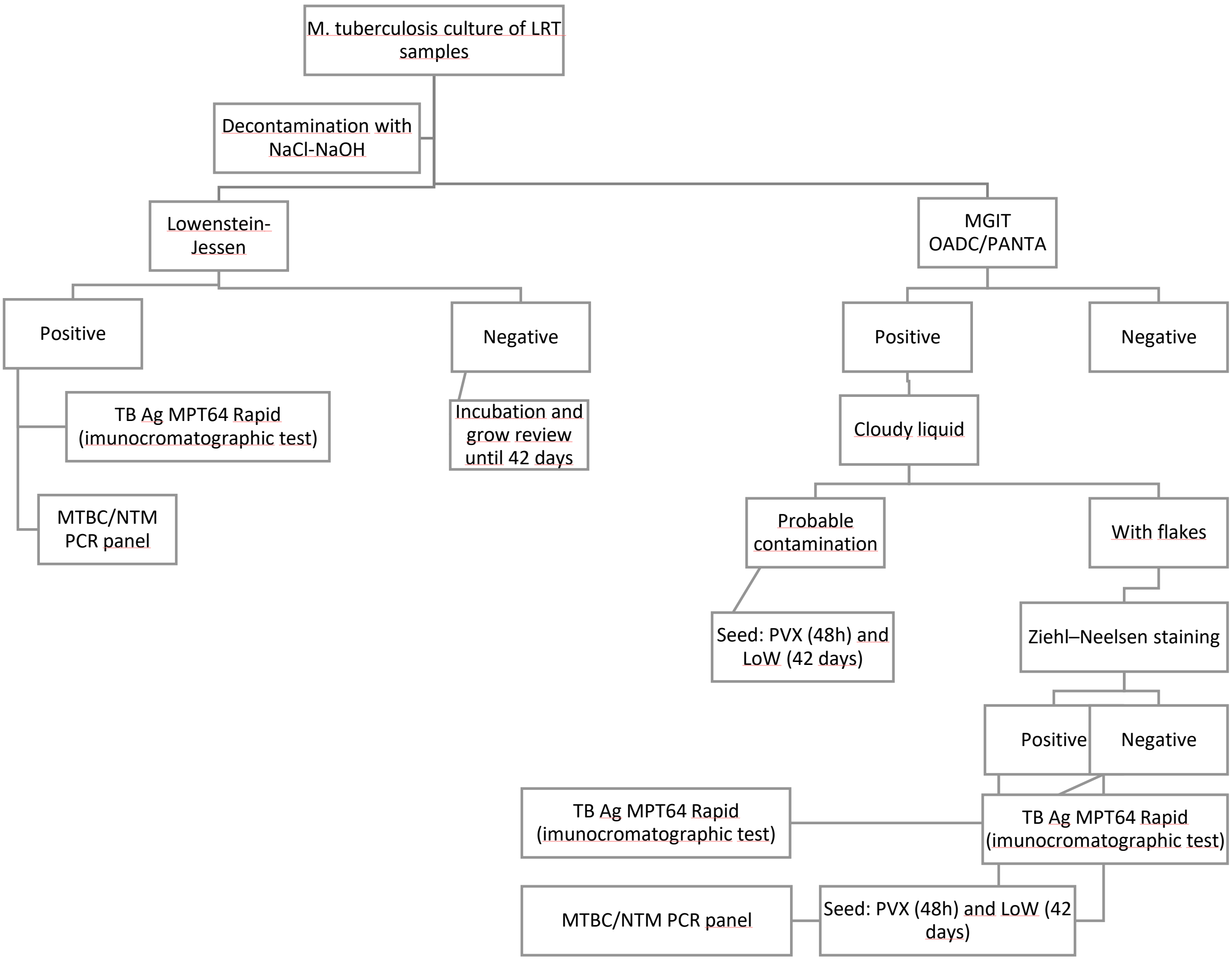


### Target-based molecular approaches for identification of etiologic agents of infection

Samples used were: nasopharyngeal swabs or BAL. Viruses panels include Influenza A (H1, H1N1 and H3) (Flu A-H1, Flu A-H1N1, Flu A-H3) and B (Flu B) viruses, Respiratory Syncytial virus A (RSV A) and B (RSV B), Adenovirus (AdV), Enterovirus (HEV), metapneumovirus (MPV), Parainfluenza virus types 1-4 (PIV1, PIV2, PIV3, PIV4), Bocavirus 1-4 (HBoV), Coronavirus 229E, NL63, OC43 and Human Rhinovirus (HRV) or *Bordetella pertussis* (BP) and *parapertussis* (BPP), *Chlamydophila pneumoniae* (CP), *Legionella pneumophila* (LP), *Mycoplasma pneumoniae* (MP), *Haemophilus influenzae* (HI) and/or *Streptococcus pneumoniae* (SP).

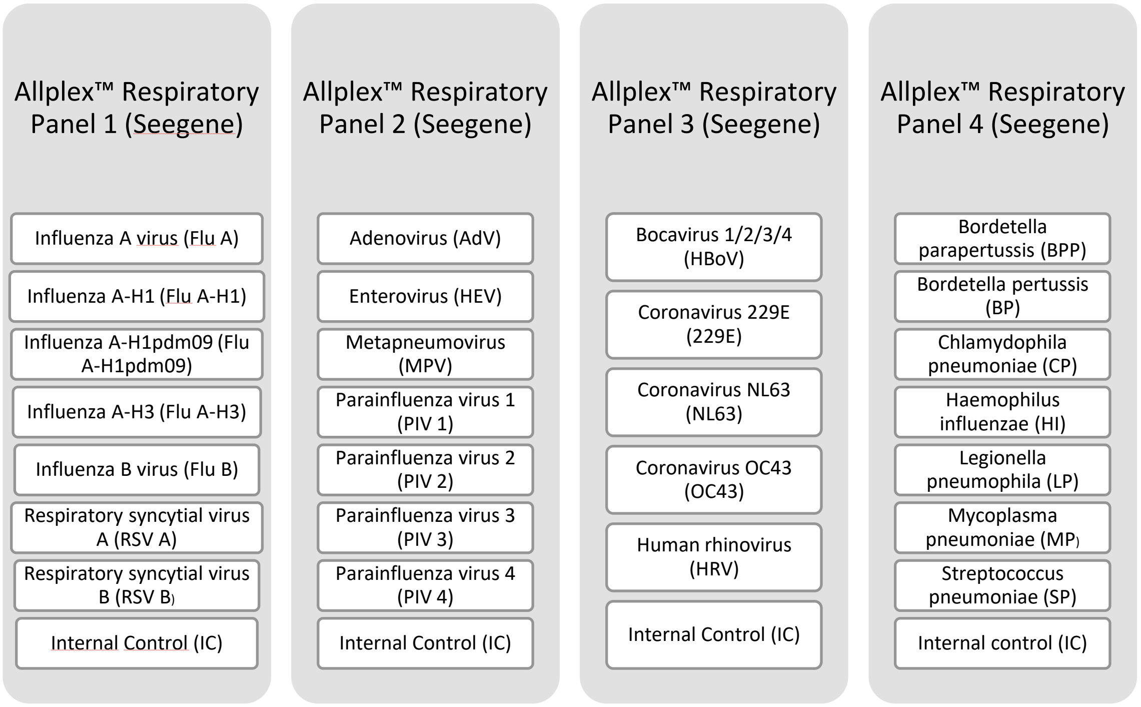


### Supplementary Tables

| Bacterial pathogens detected by molecular approach | n | % of identification |
|---|---|---|
| *Haemophilus influenzae* | 92 | 16,0% |
| *Streptococcus pneumoniae* | 31 | 5,4% |
| *Streptococcus pneumoniae and Haemophilus influenzae* | 15 | 2,6% |
| *Legionella pneumophila* | 3 | 0,5% |
| *Mycoplasma pneumoniae, Streptococcus pneumoniae and Haemophilus influenzae* | 2 | 0,3% |
| *Mycoplasma pneumoniae* | 1 | 0,2% |
| *Mycoplasma pneumoniae and Haemophilus influenzae* | 1 | 0,2% |
| *Bordetella pertussis* | 1 | 0,2% |
| *Chlamydophila pneumoniae, Streptococcus pneumoniae and Haemophilus influenzae* | 1 | 0,2% |
| *Chlamydophila pneumoniae* | 1 | 0,2% |
| *Chlamydophila pneumoniae and Haemophilus influenzae* | 1 | 0,2% |
| *Chlamydophila pneumoniae and Legionella pneumophila* | 1 | 0,2% |

**Table S1** – Number and percentage of bacterial test diagnosis obtained through the use of a target molecular approach (Allplex™ Respiratory Panel 4 (Seegene)) in BAL.

| | | |
|---|---|---|
| Top 10 | *Pseudomonas aeruginosa*<br>*Staphylococcus aureus*<br>*Haemophilus influenzae*<br>*Klebsiella pneumoniae*<br>*Escherichia coli* | *Candida albicans*<br>*Streptococcus pneumoniae*<br>*Acinetobacter baumannii*<br>*Enterobacter cloacae*<br>*Moraxella catarrhalis* |
| 54 additional species | *Enterobacter aerogenes*<br>*Proteus mirabilis*<br>*Stenotrophomonas maltophilia*<br>*Serratia marcescens*<br>*Klebsiella oxytoca*<br>*Citrobacter koseri*<br>*Morganella morganii*<br>*Neisseria meningitidis*<br>*Pasteurella multocida*<br>*Candida tropicalis*<br>*Citrobacter freundii*<br>*Corynebacterium striatum*<br>*Enterobacter cloacae cloacae*<br>*Enterococcus faecalis*<br>*Haemophilus parainfluenzae*<br>*Proteus hauseri*<br>*Streptococcus mitis*<br>*Acinetobacter bereziniae*<br>*Acinetobacter lwoffii*<br>*Bordetella bronchiseptica*<br>*Candida glabrata*<br>*Candida krusei*<br>*Corynebacterium pseudodiphtheriticum*<br>*Escherichia fergusonii*<br>*Klebsiella species*<br>*Raoultella ornithinolytica*<br>*Serratia liquefaciens* | *Serratia rubidaea*<br>*Staphylococcus epidermidis*<br>*Streptococcus oralis*<br>*Acinetobacter calcoaceticus/baumannii complex*<br>*Acinetobacter junii*<br>*Acinetobacter nosocomialis*<br>*Aeromonas sobria*<br>*Aeromonas veronii biovar sobria*<br>*Aspergillus spp.*<br>*Burkholderia cepacia*<br>*Candida dubliniensis*<br>*Citrobacter braakii*<br>*Corynebacterium propinquum*<br>*Eikenella corrodens*<br>*Enterobacter asburiae*<br>*Enterobacter bugandensis*<br>*Kocuria rosae*<br>*Morganella morganii ssp. morganii*<br>*Proteus vulgaris*<br>*Providencia rettgeri*<br>*Pseudomonas fluorescens*<br>*Pseudomonas luteola*<br>*Shewanella algae*<br>*Staphylococcus hominis*<br>*Streptococcus agalactiae - (Group B)*<br>*Streptococcus anginosus*<br>*Streptococcus gordonii* |

**Table S2** – Bacterial/fungal species isolated by culture-based methods from the different respiratory samples (suptum, ETA, BAA and BAL) and identified through MALDI-TOF. Species are ordered by frequency of isolation.

| Viral detected by molecular approach | n | % of identification |
|---|---|---|
| Influenza A virus | 14 | 5,6% |
| Rhinovirus | 6 | 2,4% |
| Influenza B virus | 2 | 0,8% |
| Metapneumovirus | 2 | 0,8% |
| Adenovirus | 1 | 0,4% |
| Adenovirus and Enterovirus | 1 | 0,4% |
| Bocavirus | 1 | 0,4% |
| Bocavirus and Rhinovirus | 1 | 0,4% |
| Coronavirus (229E) | 1 | 0,4% |
| Respiratory syncytial virus B | 1 | 0,4% |

**Table S3** - Percentage of viral positive identification through the use of Allplex™ Respiratory Panel 1-3 (Seegene) in BAL.

### Supplementary Figures

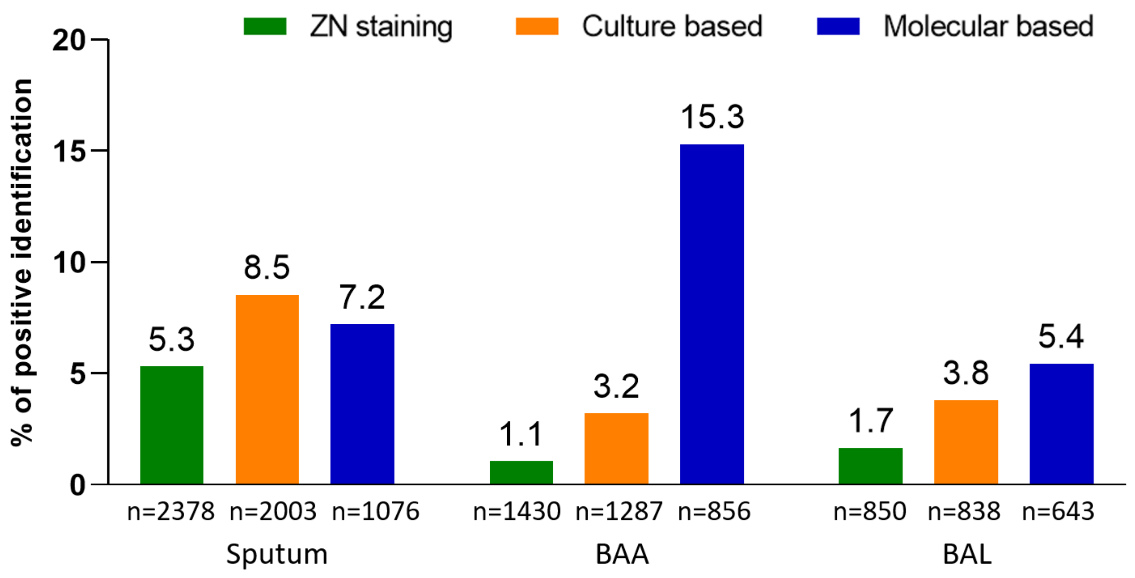


**Figure S1** – Percentage of *M. bacterium* positive identification through direct microscopic observation of Ziehl-Nielsen staining, culture-based methods and target molecular approach (MTB/NTMe - Real time detection, Anyplex™. Seegene). Positive identifications are grouped by diagnostic method.
